# Real-time air pollution and bipolar disorder symptoms: a remote monitored cross-sectional study

**DOI:** 10.1101/2022.11.21.22282067

**Authors:** Aaron Kandola, Joseph F Hayes

**Author notes:** **Corresponding author:** Joseph Hayes, 6^th^ Floor Maple House, 149 Tottenham Court Road, London.

## Abstract

Air pollution is associated with unipolar depression and other mental health outcomes. We assessed the real-time association between localised mean air quality index and the severity of depression and mania symptoms in people with bipolar disorder. We found that as air quality worsened, symptoms of depression increased. We found no association between air quality and mania symptoms.

## Background

A range of air pollutants have been found to be associated with unipolar depression in the long- and short-term, including particulate matter, nitrogen dioxide, sulphur dioxide, ozone, and carbon monoxide.^1^ However, we are only aware of one study that reports an association with bipolar symptom severity.^2^ This cross-sectional study found that increased exposure to particulate matter decreased manic episode severity, but increased the risk of mixed episodes. This finding may suggest that air pollution has depressogenic effects in bipolar disorder as well as unipolar depression.

The digital healthcare platform juli reports levels of these air pollutants to users as the daily air quality index (AQI).^3^ Here, we examine the cross-sectional association between two-week mean AQI and depression symptoms as measured by the Patient Health Questionnaire-8 (PHQ-8),^4^ and mania symptoms using the Altman Self-Rating Mania scale (ASRM)^5^ in juli users with bipolar disorder.

## Method

The digital healthcare platform juli is globally available on iOS. Users of juli consent to their aggregated de-identified data being used for research at the point of sign-up (https://www.juli.co/privacy-hub/privacy-principles). The UCL Research Ethics Committee gave ethical approval for this study (ID 19413/002). For this study, we used data from users who stated that a psychiatrist had previously diagnosed them with bipolar disorder. Users were recruited from the start of December 2020 until April 2022. All users are over the age of 18.

Our exposure was mean AQI scores over the two weeks prior to juli users completing PHQ-8 and ASRM scales. Within the platform individuals are presented with the daily AQI for their local area based on their smartphone geolocation. The AQI ranges from 0-500, with higher scores reflecting worse pollution. For each pollutant, an AQI value of 100 generally corresponds to an ambient air concentration that equals the level of the short-term national ambient air quality standard for the protection of public health. AQI values at or below 100 are satisfactory and values over 100 are unhealthy.^3^

Our outcomes were the total PHQ-8 and ASRM scores. The PHQ-8 asks eight questions about symptoms of depression over the previous two weeks and is a widely used clinical screening and research tool.^4^ The ASRM is a 5-item, multiple choice scale covering the previous week.^5^ We examined how the mean AQI over these two weeks was associated with the total score on the PHQ-8 and ASRM in individuals with bipolar disorder using linear regression. We adjusted for potential confounders of the relationship between AQI and PHQ-8/ASRM. These were: the age and gender of the user, mean daily step count, mean temperature, and mean humidity over the 14 days. As individuals could complete multiple PHQ-8 while using juli (up to two weekly), we accounted for clustering of PHQ-8 scores within individuals using robust standard errors. We performed all analyses using Python.

## Results

1,423 individuals with bipolar disorder completed 2,930 PHQ-8 and ASRM questionnaires. Of the entire cohort 1,142 (80.3%) were female and the median age was 26 (interquartile range 20-35). Of the included individuals, 701 (49.3%) had received a diagnosis for >5 years and 723 (50.8%) continue to see a psychiatrist regularly. The AQI ranged from 2-235 (mean 43, standard deviation 23). The PHQ-8 scores ranged from 0-24 (mean 12.8, standard deviation 5.9) and the ASRM scores ranged from 0-20 (mean 4.75, standard deviation 3.9).

In unadjusted models, AQI was associated with PHQ-8 scores (coefficient = 0.011, 95%CI = 0.001 to 0.022, p = 0.045). After adjusting for age, sex, mean step count, temperature and humidity, we found an association between AQI and PHQ-8 score, such that total PHQ-8 score increased by 0.011 points for every 1 point increase in AQI (95%CI 0.001 to 0.022, P=0.038). We found no association between AQI and ARSM in unadjusted (coefficient = 0.002, 95%CI -0.006 to 0.010, P=0.584) or adjusted (coefficient = 0.001, 95%CI -0.007 to 0.009, P=0.754) models. Figure 1 shows the probability of different PHQ-8 and ASRM scores for AQI <100 (healthy air) and ≥100 (unhealthy air).

**Figure 1.**
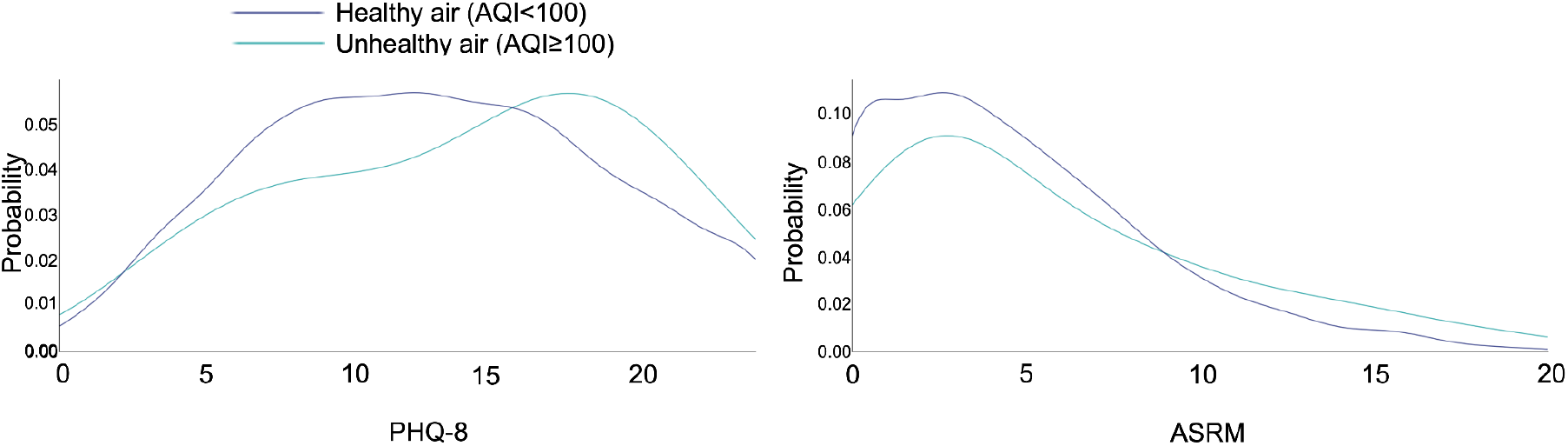
Distribution of PHQ-8 and ASRM scores by AQI

## Discussion

Ecological studies have previously found an association between air quality and rates of bipolar disorder diagnosis.^6^ This is the first study to find that real-time air pollution exposure is associated with acutely increased depression symptoms in people with bipolar disorder. The effect is small, such that one standard deviation (23 points) increase in AQI is associated with a quarter of a point increase on the PHQ-8. We did not find an association with mania symptoms as in a previous study.^2^

Our study minimises the risk of exposure misclassification that may be present in previous studies of air pollution and mental health by using the individuals mobile phone geolocation. We used a summary measure of air pollution, an approach with strengths, as most pollutants are highly correlated, but does not allow for the study of particular types of pollution.^7^ There may be unmeasured confounders that would influence our findings, particularly the effect of socioeconomic position, which juli does not capture. Included participants may not be representative of the wider population of people with bipolar disorder as they are more likely to be female, have an iPhone, and have chosen to download the juli platform.

Air pollutants potentially play a role in mental health problems through inducing neurotoxicity, neuroinflammation and hormonal dysregulation.^8^ Therefore, air pollution may represent an important modifiable risk factor for symptom severity in various mental health problems, including bipolar disorder.

## Data Availability

All data are available upon agreement with juli Health.

## Acknowledgements

JFH is supported by the UK Research and Innovation grant MR/V023373/1, the University College London Hospitals NIHR Biomedical Research Centre, and the NIHR North Thames Applied Research Collaboration. AK is supported by the UK Research and Innovation (UKRI) Digital Youth Programme award [MRC project reference MR/W002450/1], which is part of the AHRC/ESRC/MRC Adolescence, Mental Health and the Developing Mind programme.

